# Pimavanserin for the Treatment of Alzheimer’s Disease Psychosis: An Evaluation of the Clinical Importance of Efficacy Results

**DOI:** 10.1101/2022.08.11.22278482

**Authors:** Clive Ballard, Jeffrey Cummings, Pierre N Tariot, Sanjeev Pathak, Bruce Coate, Srdjan Stankovic

## Abstract

Alzheimer’s disease psychosis (ADP) is a common and serious condition with substantial unmet need for safe and effective treatments. Pimavanserin is approved in the US to treat Parkinson’s disease hallucinations and delusions. This post-hoc analysis of randomized, double-blind, placebo-controlled, phase 2 trial of nursing-home-residents with ADP evaluated the efficacy of pimavanserin by improvements (least squares mean change) in the Neuropsychiatric Inventory–Nursing Home Version Psychosis Score (NPI-NH PS). The clinical significance of the primary endpoint was assessed using responder analyses (≥ 30% and ≥ 50%); numbers needed to treat (NNT); cumulative response (0-100% improvement); All Patients and Severe Psychosis (NPI-NH PS ≥ 12) subgroups were evaluated; improvements in hallucinations and delusions by NPI-NH-PS frequency (3 or 4 points) and severity (2 or 3 points); and ≥ 50% responder analysis at earlier timepoints (weeks 2 and 4). Among 345 patients screened, 181 patients were randomized to pimavanserin (n = 90) and placebo (n = 91). Patients were elderly (mean age: 86 years) and frail (baseline mean NPI-NH PS scores of 9.5 and 10 for pimavanserin and placebo, respectively). Pimavanserin significantly improved NPI-NH PS relative to placebo (−3.76 versus −1.93, respectively; *p* = 0.0451). In responder analyses, pimavanserin demonstrated a significantly greater reduction in NPI-NH PS versus placebo at both the ≥ 30% (*p* = 0.0159) and ≥ 50% (*p* = 0.0240) thresholds with NNTs of 6 and 7, respectively. Furthermore, pimavanserin demonstrated significantly earlier reductions in NPI-NH PS compared with placebo for the ≥ 30% (*p* = 0.0336) and ≥ 50% (*p* = 0.0044) thresholds. The cumulative response analysis demonstrated significantly greater efficacy of pimavanserin for All Patients (*p =* 0.052) and Severe Psychosis (*p* = 0.004), and Severe Patients versus All Patients demonstrated a greater reduction in NPI-NH PS (p = 0.011; effect size = 0.73). Pimavanserin also demonstrated numerically greater improvements for the frequency and severity of delusions and hallucinations. Responder analyses at earlier timepoints demonstrated significantly greater response rates with pimavanserin versus placebo at week 2 (p = 0.016) but not 4 (p = 0.051). These findings support pimavanserin as safe and effective in a population of nursing-home-resident patients with ADP.

## Background

Alzheimer’s disease psychosis (ADP) is a serious and terminal condition that lacks satisfactory or US Food and Drug Administration–approved treatments. The standard of care usually includes atypical antipsychotics, despite the uncertain efficacy, lack of approval, and significant potential toxicity of these treatments for ADP.[1] The median prevalence of ADP among patients with Alzheimer’s disease dementia is 41% (with a range of 12.2%-74.1%) and a 3-year cumulative incidence of approximately 50%.[2] Furthermore, the risk for outcomes, such as nursing home admission, progression to severe dementia, and death, is approximately 1.5-2.0 times higher in dementia patients with psychosis than in those without psychosis.[3-5] Together, these findings indicate an urgent need for effective and safe ADP treatments. This unmet need for safe and effective treatments for ADP is compounded by the potentially serious safety risks of atypical antipsychotics, which include the risk of mortality, cognitive decline, and extrapyramidal symptoms.[6-8]

Pimavanserin is a selective serotonin receptor inverse agonist and antagonist at the 5-hydroxytryptamine (serotonin) receptor subtype 2A (5-HT2A) receptors and, to a lesser extent, at 5-HT2C receptors. Pimavanserin is approved for the treatment of hallucinations and delusions associated with psychosis in Parkinson’s disease in the US.[9] In a previous randomized, double-blind, placebo-controlled, 6-week, parallel-group, outpatient study of 199 patients with Parkinson’s disease psychosis, pimavanserin achieved statistically significant (*p* = 0.001) reductions in hallucinations and delusions with a least-squares mean (LSM) difference at week 6 of −3.06 (Cohen’s *d* effect size = 0.50) on the Parkinson’s disease-adapted scale for assessment of positive symptoms (SAPS-PD).[10] In addition, another study of patients with Parkinson’s disease reported that patients with cognitive impairment (Mini-Mental State Exam [MMSE] scores of 21-24) had a more robust response to pimavanserin than those with normal cognition.[11] In addition to these findings, various studies (ie, postmortem, imaging via positron emission tomography, and genetic analyses) have suggested that targeting 5-HT2A receptors could be beneficial for psychosis symptoms in Alzheimer’s disease as a well as for Parkinson’s disease.[12, 13]

As previously reported, pimavanserin was evaluated in a randomized, double-blind, placebo-controlled, phase 2 clinical trial in nursing-home-resident patients who met the criteria for ADP.[14] In this study, compared with placebo, pimavanserin achieved significantly greater reductions on the primary endpoint of the Neuropsychiatric Inventory–Nursing Home Version Psychosis Score (NPI-NH PS) at week 6.[14] Patients were monitored following the 6-week primary endpoint through week 12 to assess the safety and tolerability of pimavanserin further; the sustained efficacy of pimavanserin (beyond week 6) was evaluated in a separate study.[15]

The objective of the current study analysis was to understand the clinical importance of improvements observed over the 6-week treatment period. Specifically, in post hoc analyses, we assessed the efficacy of pimavanserin at timepoints up to and including week 6, as well as the number needed to treat analyses (NNT), an analysis of patients with severe disease, and an evaluation of improvements in hallucinations and delusions by splitting the frequency and severity ratings on the NPI-NH-PS.

## Methods

The rationale and design of the clinical trial have been described in detail in a prior publication.[14] Briefly, Study 019 (NCT02035553) was a 12-week, randomized, double-blind, placebo-controlled, phase 2, single-center study in nursing-home-resident patients (ages ≥ 50 years) from a 133-nursing home network in the United Kingdom (**Figure 1**). There was an approximate 3-week screening period in which caregivers were trained to provide psychosocial therapy using the Brief Psychosocial Therapy for Psychosis (BPST) to the patients, with a target of 5 times per week and a minimum of 3 times per week.

**Figure 1.**
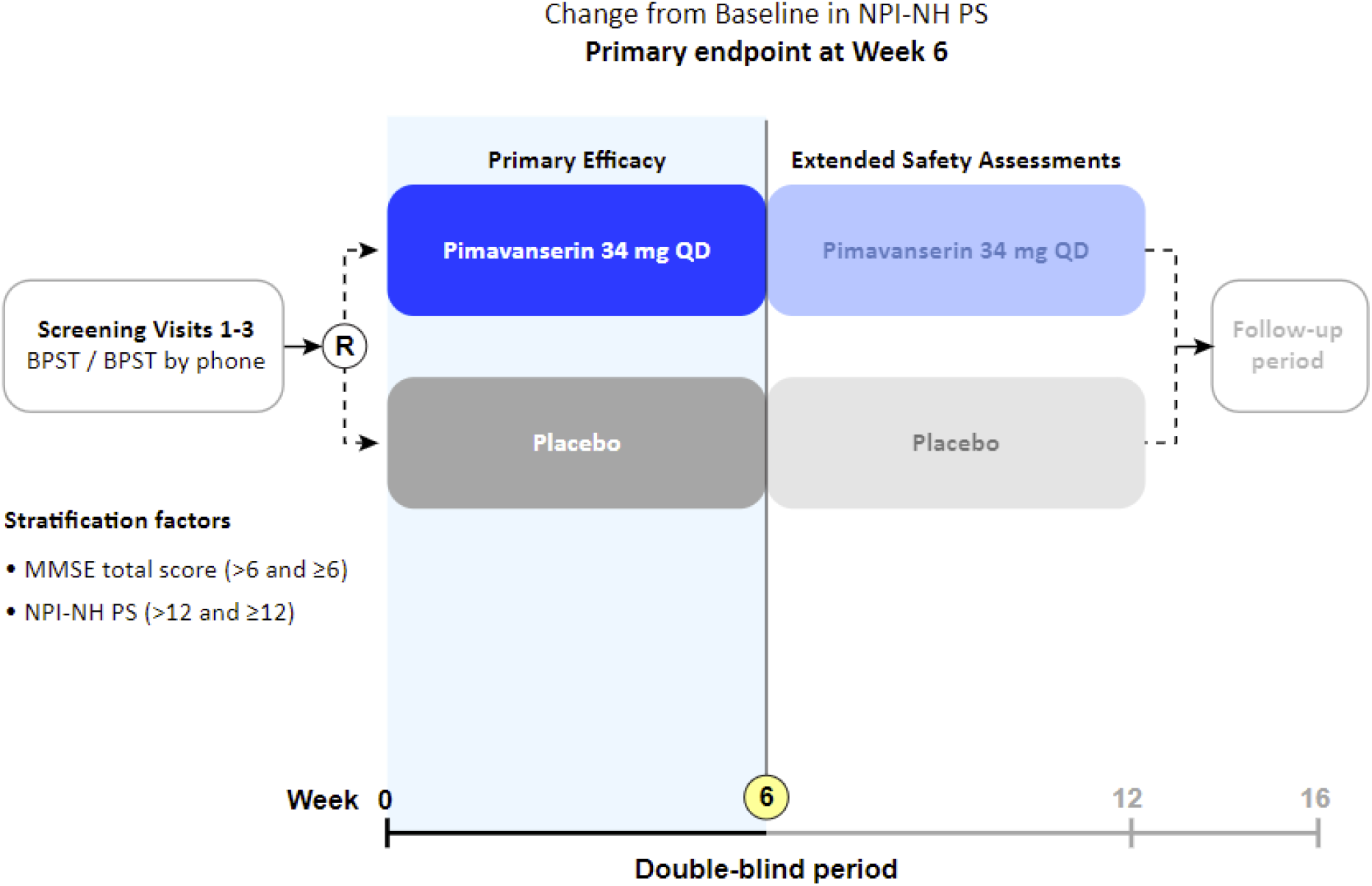
Randomized, double-blind, placebo-controlled, phase 2 design of Study 019 (NCT02035553). BPST, Brief Psychosocial Therapy for Psychosis; MMSE, Mini-Mental State Examination; NPI-NH PS, Neuropsychiatric Inventory-Nursing Home Psychosis Score; QD, every day; R, randomization.

Inclusion criteria specified that patients had to meet criteria for ADP, which included possible or probable Alzheimer’s disease as defined by the National Institute of Neurological and Communicative Disorders and Stroke–Alzheimer’s Disease and Related Disorders Association (NINCDS–ADRDA),[12] and meet the Jeste and Finkel [16] criteria for psychosis of Alzheimer’s disease. The psychotic symptoms (ie, visual or auditory hallucinations, delusions, or both) had to have developed after the diagnosis of Alzheimer’s disease. Patients were required to have a baseline score of ≥ 4 on either the hallucinations (frequency × severity) or delusions (frequency × severity) domains of the NPI-NH [17] or a total combined score of ≥ 6 (hallucinations + delusions). In addition, patients had to have a baseline MMSE score of ≥ 1 and ≤22, reside in a nursing home for ≥ 4 weeks prior to randomization, not be bedridden, and remain in the study.

### Primary Endpoint

The primary endpoint was the change from baseline to week 6 on the NPI-NH PS. The NPI-NH-PS score was chosen as the primary outcome measure; the NPI is the most common measure utilized for the assessment of neuropsychiatric symptoms in AD and has been used in many clinical studies.[17] The NPI-NH-PS includes the domains of hallucinations and delusions and is appropriate for the patient population in this study. The nursing home version (NPI-NH) of this scale was chosen as it was designed to examine psychopathology in nursing-home patients and has been validated for use in this population.[18] The score of each domain on the NPI-NH-PS, if present, represents the product of symptom frequency (range 1 to 4) and severity (range 1 to 3) for a maximum score of 12 on each domain (with higher scores denoting more serious symptoms).[17]

The week-6 timepoint on the primary endpoint was chosen based on an understanding of the natural history of psychosis in AD, which shows fluctuating symptoms often resulting in spontaneous improvement with subsequent relapse.[19-22] Therefore, to evaluate the efficacy of pimavanserin in comparison with placebo, a timepoint was chosen that was relatively early in treatment while allowing sufficient time for efficacy to be determined. The week-6 timepoint is considered the regulatory standard for antipsychotic drug development.[23, 24]

Although the primary efficacy endpoint was assessed at week 6, patients were followed in a double-blind fashion through 12 weeks to assess potential adverse impacts on cognition (as measured by MMSE). During this extended time frame, from weeks 6-12, efficacy was assessed on an exploratory basis.

### Study Conduct

The study included a dedicated group of 20 trained and experienced individuals who administered the NPI-NH, including the NPI-NH PS. Different individuals administered the assessment at consecutive visits. Those administering the NPI-NH were trained by an experienced provider of rater training and certification. There was a frequent assessment of raters to reduce drift and scoring variability. Raters were provided feedback and refresher training.

Additionally, audio recordings of care giver interviews were remotely reviewed to improve the quality of the raters on the NPI-NH. Interrater reliability was assessed for the NPI-NH via the intraclass correlation coefficients (ICC), and all values were >0.9.

### Statistical Methodology

The post hoc analyses presented here were conducted in the full analysis set to investigate the clinical importance of the primary efficacy finding. These additional analyses included analyses of ≥ 30% and ≥ 50% responders, efficacy in Severe Patients (NPI-NH PS ≥ 12 at baseline), and efficacy in patient with various frequency and severity ratings on the individual hallucination and delusion items of the psychosis score.

For the responder analysis, we evaluated the number of patients achieving ≥ 30% and ≥ 50% improvement—thresholds that represent meaningful benefit in studies of neuropsychiatric symptoms [23, 25, 26]—on the NPI-NH-PS at the primary endpoint of week 6. In patients with psychosis, there is always an urgency to improve symptoms quickly for relief to patients, caregivers, and families. Therefore, we conducted responder analyses (≥ 50% improvement) at every visit (ie, weeks 2 and week 4) prior to the primary timepoint of week 6 to evaluate the rapidity of response with pimavanserin. For this analysis, the proportion of patients meeting the definition was plotted by treatment group and visit. Time to response analyses (ie, the time of the first occurrence of a ≥ 30% or ≥ 50% reduction from baseline in NPI-NH PS) were conducted for these analyses, and the reduction had to be sustained for ≥ 7 days.

For the responder analyses, cumulative response curves on the percent change from baseline at week 6, across all possible responder cutoffs (with a range from 0 to 100% improvement), were constructed by plotting the proportion of patients meeting the cutoffs in each treatment group. To evaluate improvement from baseline (ie, responder), patients who never improved (ie, all postbaseline scores were higher than baseline) had their percent change value set to 0. In addition, the relationship between efficacy and severity of symptoms was investigated by conducting a responder (ie, ≥ 30% and ≥ 50%) analysis in All Patients and Severe Patients (NPI-NH PS ≥ 12) at the week 6 timepoint.

The NPI-NH-PS data frequency and severity scores for hallucinations and delusions was split to further contextualize these data and illustrate the benefits for patients and their caregivers. For each of the NPI-NH PS hallucinations and delusions domains, the proportion of patients improving 3 or 4 levels from baseline to week 6 on the frequency score and 2 or 3 levels from baseline to week 6 on the severity score were reported by the treatment arm. These levels of improvement represent meaningful and tangible improvements in hallucination and delusions.

For these analyses, differences in response rates between pimavanserin and placebo were analyzed using the Cochran-Mantel-Haenszel (CMH) test stratified by the baseline MMSE category (<6 and ≥ 6) and baseline NPI-NH PS category (<12 and ≥ 12). Two-sided *p*-values from a van Elteren test on the percent change from baseline, stratified by MMSE (<6 and ≥ 6) and NPI-NH PS (<12 and ≥ 12) categories, were then calculated. The numbers needed to treat (NNT) were calculated based on responder categories of a ≥ 30% or ≥ 50% improvement from baseline and included calculations for the 95% confidence intervals (CIs) [27]. Missing values were counted as nonresponders.

## Results

Patients in this study were elderly and frail, with a mean age of 86 years (**Table 1**). More than 80% of the patients were on more than 5 non-antidementia concomitant medications, reflecting many comorbid conditions typical of this population. The enrolled population is representative of a nursing-home population with severe dementia and being treated for ADP. Baseline disease characteristics and psychosis severity scores were generally consistent between groups.

**Table 1.** Demographics of enrolled ADP patients who received pimavanserin or placebo.

| **Baseline characteristics** | **Pimavanserin** | **Placebo** |
| --- | --- | --- |
|  | **N = 90** | **N = 91** |
| Age (years), mean | 86 | 86 |
| Female, % | 81 | 80 |
| White, % | 93 | 98 |
| NPI-NH PS score, mean | 9.5 | 10.0 |
| MMSE, mean | 10.2 | 9.8 |
| ≥5 non-antidementia concomitant medications, % | 82 | 85 |

ADP, Alzheimer’s disease psychosis; MMSE, Mini-Mental State Exam; NPI-NH PS, Neuropsychiatric Inventory–Nursing Home Version Psychosis Score.

A total of 345 patients were screened, and a total of 181 patients were randomized with 91 randomized to placebo and to 90 pimavanserin.

### Primary Endpoint, Efficacy at Week 6

Detailed results for the primary endpoint have been published previously and are presented only briefly here for context.[14] At week 6, pimavanserin versus placebo significantly improved NPI-NH PS (LSM change of −3.76 versus −1.93, respectively; treatment difference of −1.84; 95% CI, −3.64 to −0.04; *p* = 0.0451; and Cohen’s *d* = 0.32). At week 12, the continued improvement in the placebo group reduced the drug-placebo difference to non-significant levels.

### Clinical Importance of Efficacy Findings

#### Responder Analysis and NNTs

Differences between pimavanserin and placebo at the ≥ 30% and ≥ 50% thresholds reached nominal statistical significance (**Table 2**). At the ≥ 30% threshold, the difference between pimavanserin and placebo was 55.2% vs 37.4%, respectively (*p* = 0.0159). At the ≥ 50% threshold, the difference was 50.6% vs 34.1%, respectively (*p* = 0.0240). The NNTs for ≥ 30% and ≥ 50% thresholds were 6 (95% CI, 4-30) and 7 (95% CI, 4-46), respectively.

**Table 2.** Responder analyses and NNTs for patients with ADP treated with pimavanserin or placebo.

|  | **Pimavanserin** | **Placebo** | **Adjusted**<br>**difference** | **NNT**<br>**(95% CI)** |
| --- | --- | --- | --- | --- |
| NPI-NH PS |  |  |  |  |
| $\geq 30\%$ symptom reduction | 55.2% | 37.4% | 17% | 6 (4, 30) |
| $\geq 50\%$ symptom reduction | 50.6% | 34.1% | 16% | 7 (4, 46) |

ADP, Alzheimer's disease psychosis; NPI-NH PS, Neuropsychiatric Inventory–Nursing Home Version Psychosis Score; NNT, number needed to treat.

For the pimavanserin vs the placebo group, there was a significantly earlier occurrence of a reduction in NPI-NH PS for the ≥ 30% (*p* = 0.0336) and ≥ 50% responder group (*p* = 0.0044) (**Figure 2**).

**Figure 2.**
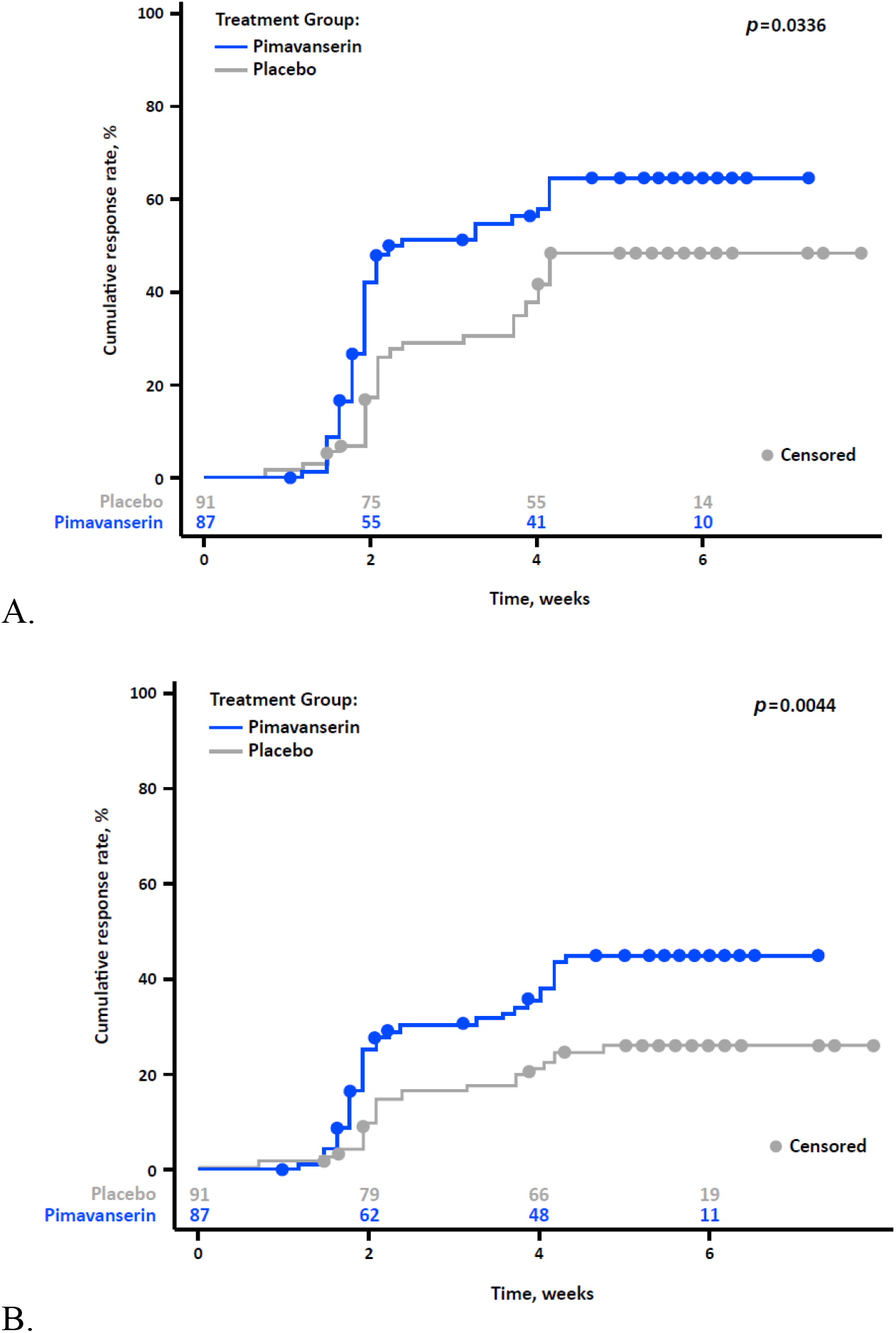
Analyses of the time to the first occurrence of a (A) ≥ 30% or (B) ≥ 50% reduction from baseline in NPI-NH PS. Analysis conducted in full analysis set, and the reduction from baseline must be confirmed for ≥ 7 days.

#### Cumulative Response Curves on Change From Baseline at Week 6

Greater efficacy with pimavanserin at week 6 was observed across the full spectrum of improvement (0%-100% improvement); nominal *p* values from the nonparametric (van Elteren) test of the 2 curves for All Patients and Severe Psychosis were 0.052 and 0.004, respectively.

NPI-NH PS, Neuropsychiatric Inventory–Nursing Home Version Psychosis Score.

#### Severe Patients And All Patients

In the prespecified subgroup of Severe Patients, the magnitude of the reduction in NPI-NH PS at Week 6 was larger than for the All Patients group (*p* = 0.011; effect size = 0.73). For All Patients, when the response was defined as a ≥ 30% reduction, the pimavanserin vs the placebo response was 55% vs 37%, respectively (*p* = 0.0159), and when defined as a ≥ 50% reduction was 51 vs 34%, respectively (*p* = 0.0240; **Figure 3**). For All Patients, the NNTs for the ≥ 30% and ≥ 50% reduction were 6 and 7, respectively.

**Figure 3.**
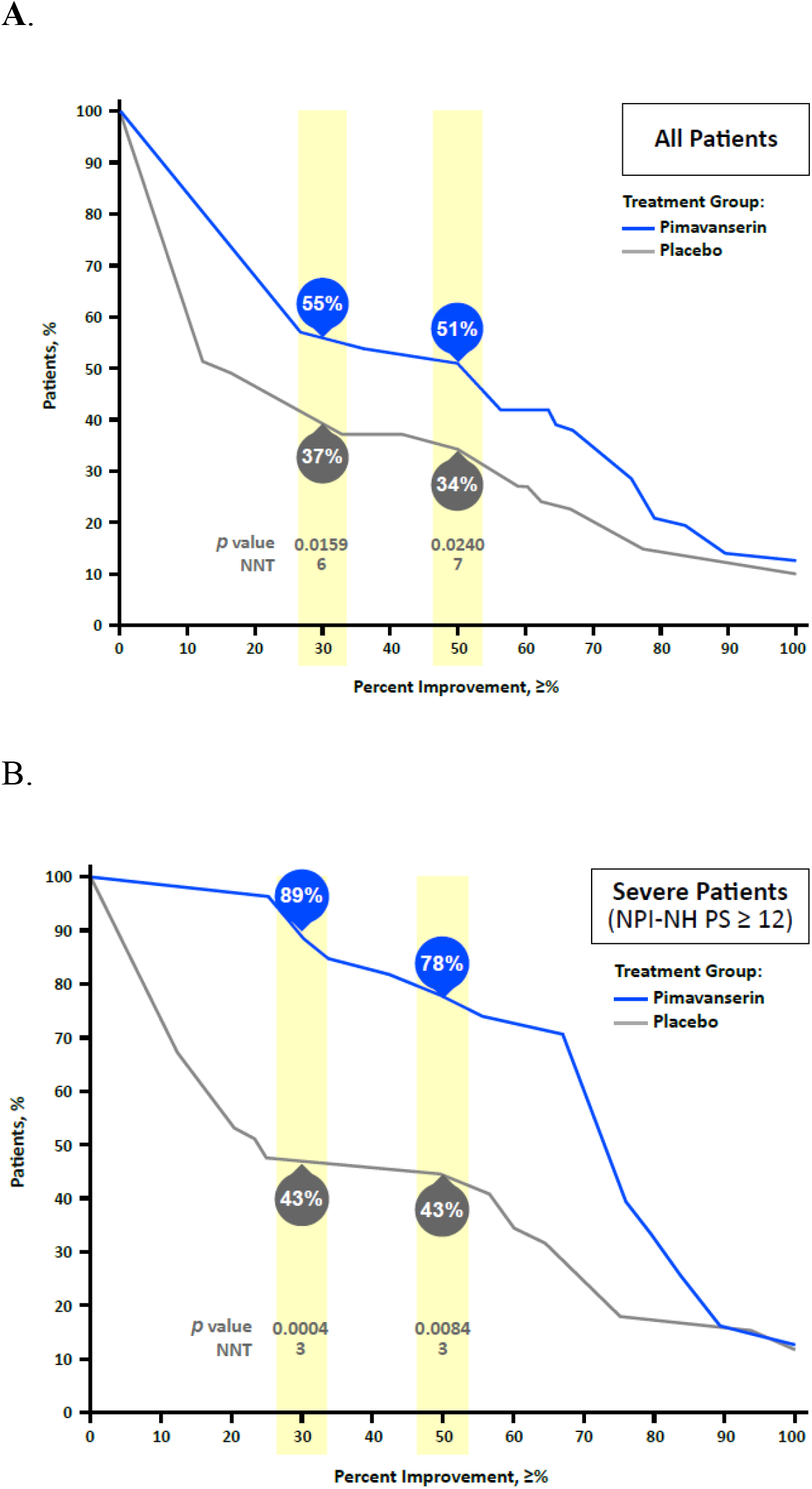
Responder Analysis of (A) All Patients and (B) Severe Patients at the Week 6 Time Point. NPI-NH PS, Neuropsychiatric Inventory–Nursing Home Version Psychosis Score; NNT, number needed to treat.

For Severe Patients, when the response was defined as a ≥ 30% reduction, the pimavanserin vs the placebo response was 88.9% vs 43.3%, respectively (*p* = 0.0004), and when defined as a ≥ 50% reduction was 77.8% vs 43.3%, respectively (*p* = 0.0084; **Figure 3**). For Severe Patients, the NNTs for the ≥ 30% and ≥ 50% reduction were both 3.

#### Frequency and Severity of Hallucinations and Delusions

For the analysis of frequency and severity of the NPI-NH-PS symptom of delusions, approximately 25% of the patients treated with pimavanserin improved by 3 or 4 points in frequency (representing a change from multiple times a day to none or less than once a week) vs 16% for the placebo group (**Figure 4**). In addition, 32% treated with pimavanserin improved by 2 or 3 points on severity (representing a change from moderate or severe distress to no or mild distress) vs 19% for placebo.

**Figure 4.**
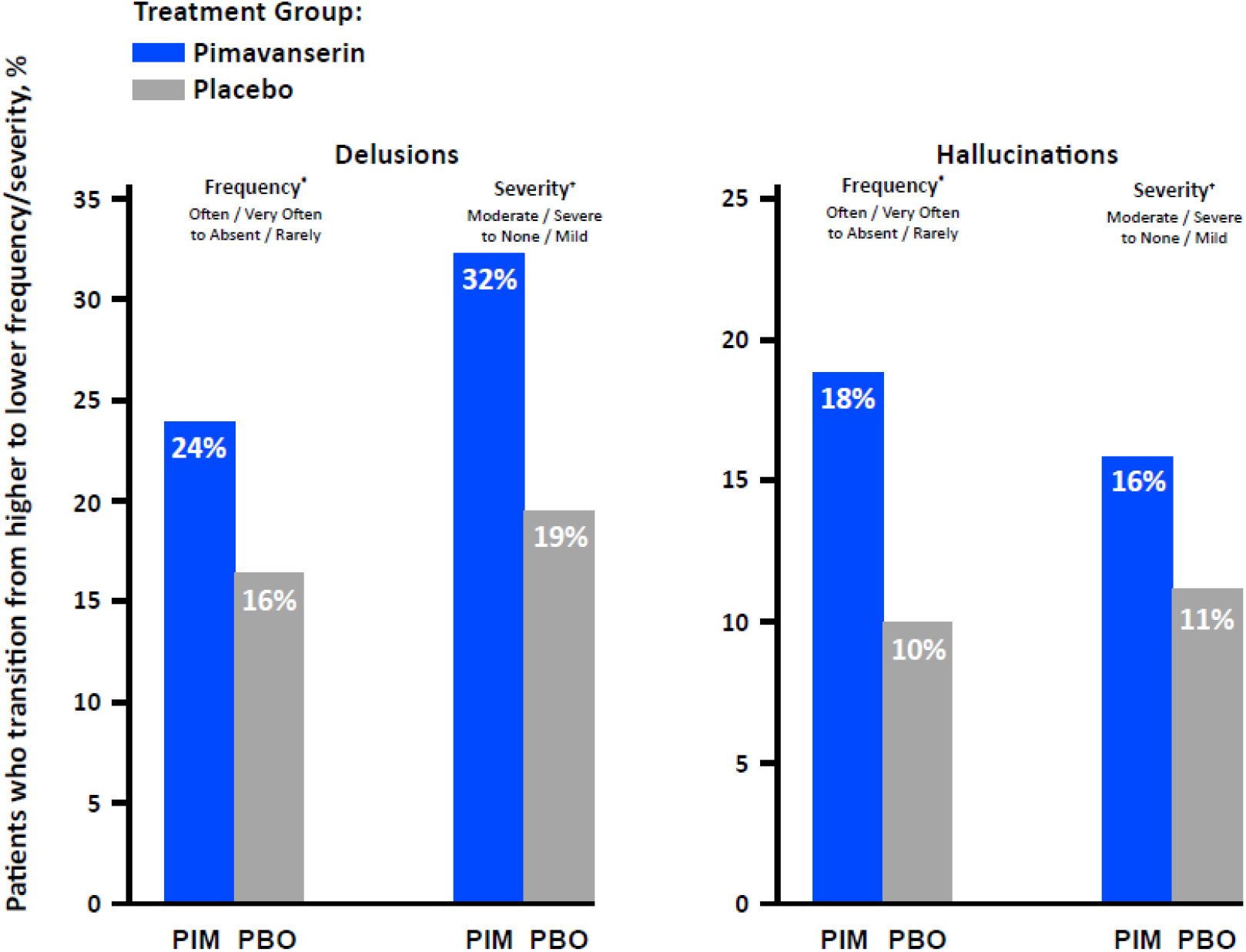
Patients Who Transition From Higher to Lower Frequency/Severity of NPI-NH-PS Symptoms of delusions and hallucinations ^*^Frequency scores were improved by 3 or 4 points, which corresponds to a transition form often/very often to absent/rarely. *Severity scores were improved by 2 or 3 points, which corresponds to a transition form moderate/severe to none/mild. PBO, placebo; PIM, pimavanserin.

Fewer patients experienced hallucinations than delusions at baseline, and hallucinations were generally less severe than delusions. In these small samples, for pimavanserin there was a nonsignificant trend for improvement in both the frequency and the severity of hallucinations (18% on frequency and 16% on severity), which has numerically higher than for placebo.

#### Responder Analysis (≥ 50% improvement) at Early Timepoints of Weeks 2 and 4

For the responder analyses (≥ 50% improvement) at the visits prior to the primary timepoint, the response rates were numerically greater than placebo, and the differences were statistically significant at week 2 (*p* = 0.016; **Figure 5**).

**Figure 5.**
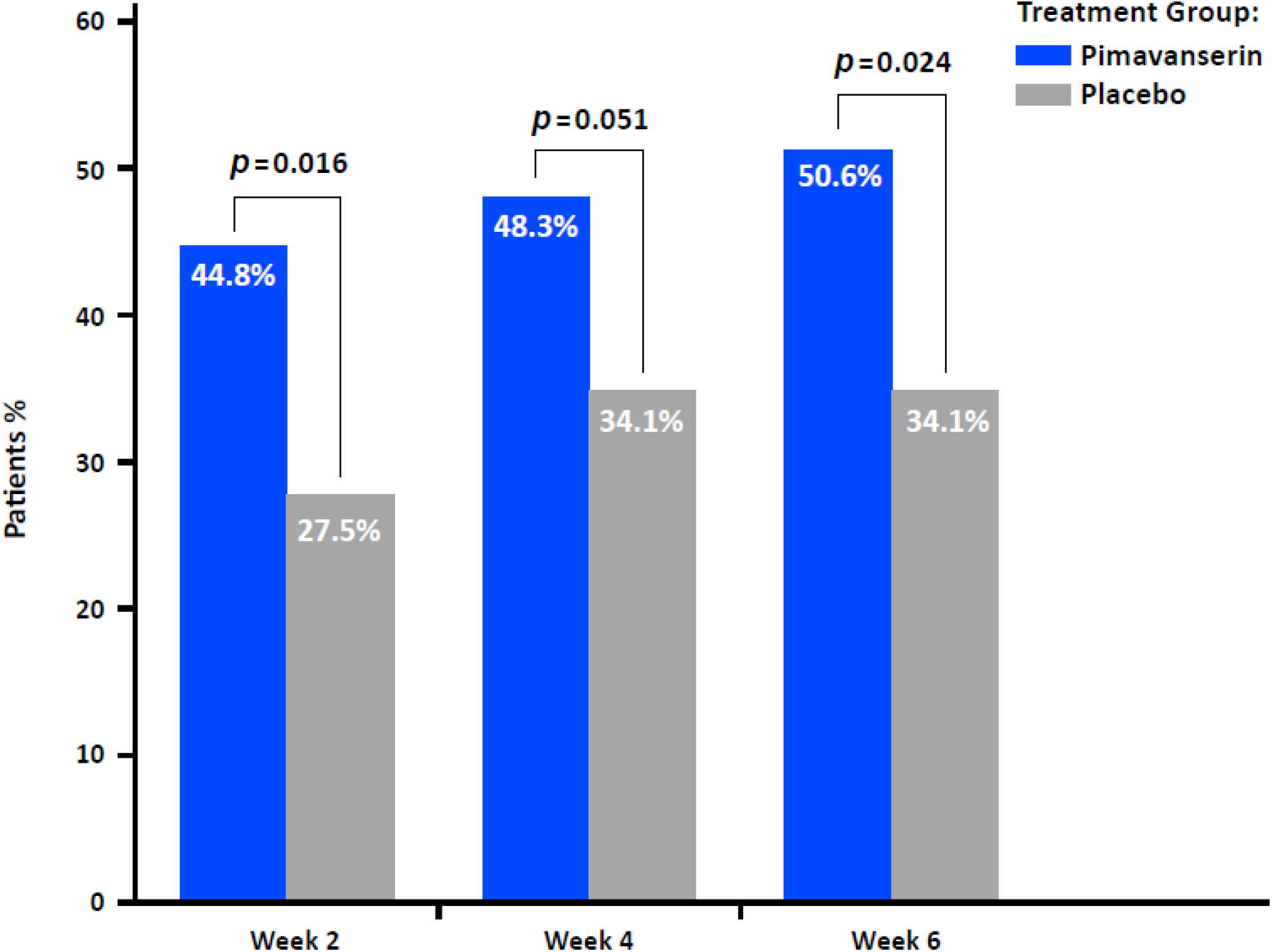
NPI-NH PS ≥ 50% Responders Across Visits at Weeks 2, 4, and 6 (Primary Efficacy Endpoint)

### Safety

The safety and tolerability of pimavanserin in this study were consistent with the well-characterized profile of pimavanserin in neurodegenerative conditions, as previously described.[14] Briefly, the most common adverse events were falls, urinary tract infections, and agitation. The frequencies of falls and urinary tract infections were similar between treatment groups. Consistent with previous reports, QT interval prolongation corrected by the Fridericia method (QTcF) was observed among patients receiving pimavanserin. There were no observed differences in mortality rates between the pimavanserin- and placebo-treated groups in this study. Over the 12-week period, there were no adverse effects on cognition between the pimavanserin- and placebo-treated groups, as assessed by the MMSE total score. Similarly, the mean Unified Parkinson’s Disease Rating Scale part III from baseline to week 12 were comparable in both treatment groups, indicating no adverse effects on motor function following treatment with pimavanserin.

## Discussion

Study 019 was a parallel-group, double-blind, placebo-controlled study that demonstrated statistically significant efficacy of pimavanserin 34 mg for the treatment of psychosis in patients with AD.[14] In this post-hoc analysis of NPI-NH PS scores, we report the efficacy of pimavanserin at timepoints up to and including week 6 (primary endpoint). The efficacy was clinically consistent, as demonstrated by analyses of responders, patients with severe disease, and the frequency and severity of hallucinations and delusions, and clinically meaningful as demonstrated by the NNT analyses. Responder analyses demonstrated that pimavanserin versus placebo demonstrated greater reductions in NPI-NH PS at all cutoffs. In addition, Severe Patients with severe psychosis (NPI-NH PS ≥ 12 at baseline) vs the All Patients group demonstrated greater reductions in NPI-NH PS, which is clinically important as patients with severe psychosis are in the greatest need of pharmacologic therapy. Together, these efficacy findings are clinically relevant in demonstrating responses of a magnitude and time to onset that are important for patients and caregivers of patients with ADP.

We evaluated the NNTs and analyses of the frequency and severity scores for hallucinations and delusions domains separately. The NNTs of 6 and 7 reported for the ≥ 30% and ≥ 50% responder groups are consistent with NNTs of <10, which have been deemed clinically meaningful [28]. The analyses of the frequency and severity of hallucinations and delusions demonstrated reductions in both dimensions of psychosis with frequency transitions from often/very often to absent/rarely and severity changes from moderate/severe to none/mild.

The key weakness of this study is the post-hoc nature of the analyses,. The strengths of the study include generalizability of the enrolled patients to a large patient population, the high inter-rater reliability of the raters (ICC > 0.9), and the use of valid and well understood scales. The generalizability of the study population is demonstrated by the fact that our population was of an advanced age, taking multiple non-antidementia concomitant medications and is representative of a nursing-home population with severe dementia with ADP. Furthermore, the reliability of the scales utilized are demonstrated by the NPI-NH’s high internal consistency (α = 0.67)[29] and high test-retest reliability as demonstrated by internal consistency values for delusions (0.89, 95% CI 0.79-0.94) and hallucinations (0.74, 95% CI 0.51-0.86).[30]. In addition, there is strong convergent validity between the NPI-NH Psychosis Factor and the Scale for Nursing Assessment in Geriatric Psychiatry psychotic features (*r* = 0.54).[29].

## Conclusions

In summary, pimavanserin demonstrated efficacy in a population of patients with ADP, who are residents in nursing homes. The efficacy was statistically significant in drug-placebo comparisons, had a robust magnitude, and had an early onset. Efficacy was apparent in those with more severe ADP as well as in the total population. No new safety or tolerability issues were observed in the study. Pimavanserin is approved for psychosis in Parkinson’s disease and this study supports its efficacy in patients with ADP.

## Data Availability

All data produced in the present work are contained in the manuscript

## References

1. Maher, A.R., et al., Efficacy and comparative effectiveness of atypical antipsychotic medications for off-label uses in adults: a systematic review and meta-analysis. Jama, 2011. 306(12): p. 1359–1369.

2. Murray, P.S., et al., Psychosis in Alzheimer’s disease. Biological psychiatry, 2014. 75(7): p. 542–552.

3. Peters, M.E., et al., Neuropsychiatric symptoms as predictors of progression to severe Alzheimer’s dementia and death: the Cache County Dementia Progression Study. American Journal of Psychiatry, 2015. 172(5): p. 460–465.

4. Rashid, N., et al., Real-World Medication Treatment Patterns for Long-Term Care Residents with Dementia-Related Psychosis. Gerontology and Geriatric Medicine, 2021. 7: p. 23337214211016565.

5. Scarmeas, N., et al., Delusions and hallucinations are associated with worse outcome in Alzheimer disease. Archives of neurology, 2005. 62(10): p. 1601–1608.

6. Vigen, C.L., et al., Cognitive effects of atypical antipsychotic medications in patients with Alzheimer’s disease: outcomes from CATIE-AD. Am J Psychiatry, 2011. 168(8): p. 831–9.

7. Ballard, C., et al., Quetiapine and rivastigmine and cognitive decline in Alzheimer’s disease: randomised double blind placebo controlled trial. BMJ, 2005. 330(7496): p. 874.

8. Schneider, L.S., K. Dagerman, and P.S. Insel, Efficacy and adverse effects of atypical antipsychotics for dementia: meta-analysis of randomized, placebo-controlled trials. The American Journal of Geriatric Psychiatry, 2006. 14(3): p. 191–210.

9. Hacksell, U., et al., On the discovery and development of pimavanserin: a novel drug candidate for Parkinson’s psychosis. Neurochem Res, 2014. 39(10): p. 2008–17.

10. Cummings, J., et al., Pimavanserin for patients with Parkinson’s disease psychosis: a randomised, placebo-controlled phase 3 trial. Lancet, 2014. 383(9916): p. 533–40.

11. Espay, A.J., et al., Pimavanserin for Parkinson’s Disease psychosis: Effects stratified by baseline cognition and use of cognitive-enhancing medications. Mov Disord, 2018. 33(11): p. 1769–1776.

12. McKhann, G., Report of the NINCDS-ADRDA work group under the auspices of department of health and human service task force on Alzheimer’s disease. Neurology, 1984. 34: p. 939–944.

13. Nordstrom, A.-L., et al., PET analysis of the 5-HT2A receptor inverse agonist ACP-103 in human brain. International Journal of Neuropsychopharmacology, 2008. 11(2): p. 163–171.

14. Ballard, C., et al., Evaluation of the safety, tolerability, and efficacy of pimavanserin versus placebo in patients with Alzheimer’s disease psychosis: a phase 2, randomised, placebo-controlled, double-blind study. The Lancet Neurology, 2018. 17(3): p. 213–222.

15. Tariot, P.N., et al., Trial of Pimavanserin in Dementia-Related Psychosis. N Engl J Med, 2021. 385(4): p. 309–319.

16. Jeste, D.V. and S.I. Finkel, Psychosis of Alzheimer’s disease and related dementias: diagnostic criteria for a distinct syndrome. The American Journal of Geriatric Psychiatry, 2000. 8(1): p. 29–34.

17. Cummings, J.L., et al., The Neuropsychiatric Inventory: comprehensive assessment of psychopathology in dementia. Neurology, 1994. 44(12): p. 2308–2308.

18. Wood, S., et al., The use of the neuropsychiatric inventory in nursing home residents: characterization and measurement. The American Journal of Geriatric Psychiatry, 2000. 8(1): p. 75–83.

19. Ballard, C.G., et al., Paranoid features in the elderly with dementia. International Journal of Geriatric Psychiatry, 1991. 6(3): p. 155–157.

20. Ballard, C.G., et al., The prevalence and phenomenology of psychotic symptoms in dementia sufferers. International Journal of Geriatric Psychiatry, 1995. 10(6): p. 477–485.

21. De Deyn, P., et al., Aripiprazole for the treatment of psychosis in patients with Alzheimer’s disease: a randomized, placebo-controlled study. Journal of clinical psychopharmacology, 2005. 25(5): p. 463–467.

22. Katz, I., et al., The efficacy and safety of risperidone in the treatment of psychosis of Alzheimer’s disease and mixed dementia: a meta-analysis of 4 placebo-controlled clinical trials. International Journal of Geriatric Psychiatry: A journal of the psychiatry of late life and allied sciences, 2007. 22(5): p. 475–484.

23. EMA, European Medicines Agency. Guideline on clinical investigation of medicinal products, including depot preparations in the treatment of schizophrenia. https://www.ema.europa.eu/en/documents/scientific-guideline/guideline-clinical-investigation-medicinal-products-including-depot-preparations-treatment_en.pdf; 2012 [accessed 27, August 2021].

24. Younis, I.R., et al., Association of End Point Definition and Randomized Clinical Trial Duration in Clinical Trials of Schizophrenia Medications. JAMA Psychiatry, 2020. 77(10): p. 1064–1071.

25. Durgam, S., et al., Cariprazine in acute exacerbation of schizophrenia: a fixed-dose, phase 3, randomized, double-blind, placebo-and active-controlled trial. The Journal of clinical psychiatry, 2015. 76(12): p. 2310.

26. Leucht, S., Measurements of response, remission, and recovery in schizophrenia and examples for their clinical application. The Journal of clinical psychiatry, 2014. 75(suppl 1): p. 11378.

27. Citrome, L., Adjunctive aripiprazole, olanzapine, or quetiapine for major depressive disorder: an analysis of number needed to treat, number needed to harm, and likelihood to be helped or harmed. Postgraduate medicine, 2010. 122(4): p. 39–48.

28. Citrome, L. and T. Ketter, When does a difference make a difference? Interpretation of number needed to treat, number needed to harm, and likelihood to be helped or harmed. International journal of clinical practice, 2013. 67(5): p. 407–411.

29. Lange, R.T., G.A. Hopp, and N. Kang, Psychometric properties and factor structure of the Neuropsychiatric Inventory Nursing Home version in an elderly neuropsychiatric population. International journal of geriatric psychiatry, 2004. 19(5): p. 440–448.

30. Chen, S., et al., Reliability and structural validity of the C hinese version of the N europsychiatric I nventory, N ursing H ome version. Psychogeriatrics, 2018. 18(2): p. 113–122.

